# Extent of cord pathology in the lumbosacral enlargement in non-traumatic versus traumatic spinal cord injury

**DOI:** 10.1101/2021.10.16.21264514

**Authors:** Gergely David, Kevin Vallotton, Markus Hupp, Armin Curt, Patrick Freund, Maryam Seif

## Abstract

**Objectives:** This study compares remote neurodegenerative changes caudal to a cervical injury in degenerative cervical myelopathy (DCM) (i.e. non-traumatic) and incomplete traumatic spinal cord injury (tSCI) patients, using MRI-based tissue area measurements and diffusion tensor imaging (DTI).

**Methods:** Eighteen mild to moderate DCM patients with sensory impairments (mean mJOA score: 16.2), 14 incomplete tetraplegic tSCI patients (AIS C&D), and 20 healthy controls were recruited. All participants received DTI and T2*-weighted scans in the lumbosacral enlargement (caudal to injury) and at C2/C3 (rostral to injury). MRI readouts included DTI metrics in the white matter (WM) columns and cross-sectional WM and gray matter area. One-way ANOVA with Tukey post-hoc comparison (p<0.05) was used to assess group differences.

**Results:** In the lumbosacral enlargement, compared to DCM, tSCI patients exhibited decreased fractional anisotropy in the lateral (tSCI vs. DCM, −11.9%, p=0.007) and ventral WM column (−8.0%, p=0.021), and showed trend toward lower values in the dorsal column (−8.9%, p=0.068). At C2/C3, no differences in DTI metrics were observed between DCM and tSCI, but compared to controls, fractional anisotropy was lower in both groups in the dorsal (DCM vs. controls, −7.9%, p=0.024; tSCI vs. controls, −10.0%, p=0.007) and in the lateral column (DCM: −6.2%, p=0.039; tSCI: −13.3%, p<0.001). WM areas were not different between patient groups, but were significantly lower compared to healthy controls both in the lumbosacral enlargement (DCM: −16.9%, p<0.001; tSCI, −10.5%, p=0.043) and at C2/C3 (DCM: −16.0%, p<0.001; tSCI: −18.1%, p<0.001).

**Conclusion:** In conclusion, mild to moderate DCM and incomplete tSCI lead to similar degree of degeneration of the dorsal and lateral columns at C2/C3, but tSCI results in more widespread white matter damage in the lumbosacral enlargement. These remote changes are likely to contribute to the impairment and recovery of the patients. Diffusion MRI is a sensitive tool to assess remote pathological changes in DCM and tSCI patients.

## Introduction

Degenerative cervical myelopathy (DCM), the most common form of non-traumatic spinal cord injury, involves a chronic mechanical compression of the cervical spinal cord.^1,2^ Traumatic spinal cord injury (tSCI) results from an immediate mechanical insult to the spinal cord leading to sensorimotor deficits and autonomic dysfunction.^3^ While DCM can stay asymptomatic for a long time (months to years)^4^, the accumulation of degenerative changes often results in progressive motor and sensory impairments, although usually less severe than in tSCI.^5,6^ Animal models demonstrated several common pathophysiological features at the injury site including necrosis^7,8^, apoptosis^9^, axonal degeneration^10^, demyelination^9,11,12^, neuroinflammation^9,13^, and changes in microvasculature^14^. In both diseases, the pathological changes are not restricted to the injury site, but spread rostrally and caudally via anterograde, retrograde, and trans-synaptic degeneration,^2,6,15–17^ although the reach of these neurodegenerative mechanisms is not fully clear. However, there are also fundamental differences in the onset, progression, and nature of secondary injury mechanisms between DCM and tSCI (abrupt onset in traumatic SCI vs. slowly developing symptoms in DCM).^6^ Consequently, it is not clear whether DCM and tSCI patients with similar functional deficits present with similar magnitudes of remote micro- and macrostructural degenerative changes.

Diffusion tensor imaging (DTI) is a magnetic resonance imaging (MRI) technique to probe white matter integrity, and is especially useful to characterize the secondary degeneration remote to the injury site, where conventional MRI technologies often fail to detect abnormality.^18–20^ Metrics derived from DTI have also been shown to correlate with disease severity both in DCM^21–25^ and tSCI^26–28^, providing means to assess the grade of microstructural damage to white matter. By applying DTI in vivo at C2/C3 (rostral to injury), Seif et al. recently showed that DCM patients feature less altered DTI indices compared to tSCI patients, but largely similar values when comparing only to incomplete tSCI patients.^29^ Although DCM and tSCI have already been compared to healthy controls, a cross-comparison between these two aetiologies has not been performed caudal to the cervical myelopathy such as the lumbosacral enlargement, a region crucial for lower motor control^30^, bladder^31^ and sexual functions^32^.

In this study we explore similarities and differences in the radiological presentation of DCM and incomplete tSCI with cervical injury in the lumbosacral enlargement, both in terms of microstructural changes and tissue atrophy, and compare these findings to those reported in the upper cervical cord. To this end, we utilize spinal cord DTI to characterize the tissue integrity in the white matter columns and high-resolution T2*-weighted structural imaging to measure the cross-sectional area of gray and white matter.

## Materials and Methods

### Study participants

Eighteen non-traumatic (DCM) patients (7 females, age (mean±std): 55.4±9.7 years), 14 tSCI patients (no female, 48.0±12.9 years), and 20 healthy controls (4 females, 42.7±15.3 years) participated in the study (Table 1). Inclusion criteria were (i) no pre-existing neurological or mental disorders, (ii) no previous spine operations, (iii) no history of head and brain lesions, (iv) no MRI contraindications, (v) >18 years of age, (vi) for DCM patients, observed spinal canal stenosis in MRI and clinical diagnosis of DCM^33^, (vii) for tSCI patients, chronic stage of injury (>12 months), incomplete injury (AIS C-E), and tetraplegic injury. tSCI patients were scanned on average (±std) 5.7±4.9 years after injury. The study protocol was designed in accordance with the Declaration of Helsinki and was approved by the local ethics committee (Kantonale Ethikkommission Zürich, EK-2010-0271). All participants provided written informed consent prior to study enrolment. Previous studies included parts of the DCM^34^, tSCI^28^, and healthy cohort^28,34^.

**Table 1.** Demographic and clinical Information of the DCM patients.

| ID | Sex | Age (yrs) | Stenosis level | Nurick score | mJOA score | AIS | UEMS score | LEMS score | LT score | PP score | T2-hyperintensity (myelopathy) |
| --- | --- | --- | --- | --- | --- | --- | --- | --- | --- | --- | --- |
| 1 | F | 51-55 | C5/6* | 1 | 17 | D | 50 | 50 | 108 | 107 | no |
| 2 | F | 66-70 | C5/6* | 1 | 16 | D | 50 | 50 | 112 | 110 | yes |
| 3 | M | 51-55 | C5/6* | 1 | 15 | D | 50 | 50 | 112 | 110 | no |
| 4 | M | 56-60 | C6/7* | 0 | 17 | D | 50 | 50 | 103 | 104 | no |
| 5 | F | 46-50 | C5/6 | 0 | 18 | E | 50 | 50 | 112 | 112 | no |
| 6 | F | 41-45 | C5/6* | 1 | 17 | D | 45 | 50 | 112 | 111 | no |
| 7 | F | 31-35 | C5/6 | 2 | 14 | D | 50 | 50 | 98 | 103 | yes |
| 8 | M | 51-55 | C5/6 | 0 | 18 | D | 49 | 50 | 90 | 90 | no |
| 9 | M | 56-60 | C5/6 | 0 | 18 | E | 50 | 50 | 112 | 112 | no |
| 10 | M | 56-60 | C6/7* | 1 | 16 | D | 50 | 50 | 111 | 108 | no |
| 11 | M | 66-70 | C5/6 | 1 | 17 | D | 50 | 50 | 112 | 110 | no |
| 12 | F | 51-55 | C5/6 | 1 | 12 | C | 50 | 50 | 84 | 84 | no |
| 13 | M | 51-55 | C5/6* | 1 | 16 | D | 50 | 50 | 110 | 107 | yes |
| 14 | M | 56-60 | C5/6* | 2 | 14 | D | 50 | 50 | 109 | 109 | yes |
| 15 | M | 61-65 | C4/5 | 0 | 18 | E | 50 | 50 | 112 | 112 | yes |
| 16 | F | 66-70 | C4/5* | 0 | 18 | E | 50 | 50 | 112 | 112 | no |
| 17 | M | 71-75 | C5/6* | 0 | 18 | E | 50 | 50 | 112 | 112 | yes |
| 18 | M | 41-45 | C5/6 | 1 | 13 | D | 49 | 50 | 85 | 86 | no |

AIS = American Spinal Injury Association Impairment Scale; mJOA = modified Japanese Orthopedic Association score (max. 18); UEMS = International Standards for Neurologic Classification of Spinal Cord Injury (ISNCSCI) upper extremity motor score (max. 50); LEMS = ISNCSCI lower extremity motor score (max. 50); LT = ISNCSCI light touch score (max: 112); PP = ISNCSCI pinprick score (max: 112). \* Multi-segmental cervical spine stenosis.

### MRI acquisition protocol

MRI scanning was performed on a 3T Siemens Skyra^Fit^ scanner, using a radiofrequency transmit coil for transmission and a 16-channel head and neck coil and a spine matrix coil for reception. An MRI compatible cervical collar was used to reduce involuntary neck motion.^35^Foam wedges were placed under the knees to minimize the distance between the lower spine and the spine matrix coil.^36^ The imaged regions included the (i) lumbosacral spinal cord (SC) with the field of view (FOV) centered at the lumbosacral enlargement and the (ii) upper cervical cord with the FOV centered at the C2/C3 disc. Slice prescription was done on a sagittal T2-weighted 2D turbo spin echo image of the cervical and lumbosacral cord, respectively (Fig. 1), where the lumbosacral enlargement was identified as the widest point of the lumbosacral cord as appearing on the T2-weighted image. In both regions, axial T2*-weighted images were acquired using a 3D multi-echo spoiled gradient-echo image sequence (Siemens MEDIC, Fig. 1) with the following parameters: 20 slices with slice thickness=2.5 mm, in-plane resolution=0.5×0.5 mm^2^, FOV=192×162 mm^2^, TE=19 ms, TR=44 ms, flip angle=11°, readout bandwidth=260 Hz/pixel, 4 repetitions, total acquisition time=7:16 min. Diffusion-weighted images for diffusion tensor imaging (DTI) were acquired using a reduced-FOV single-shot spin-echo EPI sequence consisting of 60 diffusion-weighted (b=500 s/mm^2^) and 7 T2-weighted (b=0 s/mm^2^) images. Slice prescription was identical to the MEDIC sequence and acquisition parameters were: slice thickness 5 mm (+10% gap), in-plane resolution=0.76×0.76 mm^2^, FOV=133×30 mm^2^, TE=73 ms, TR=350 ms, acquisition time around 8 min depending on the heart rate. Cardiac gating was used to reduce pulsation artifacts and two saturation bands were placed anterior and posterior of the cord to avoid fold-over artifacts.

**Fig. 1.**
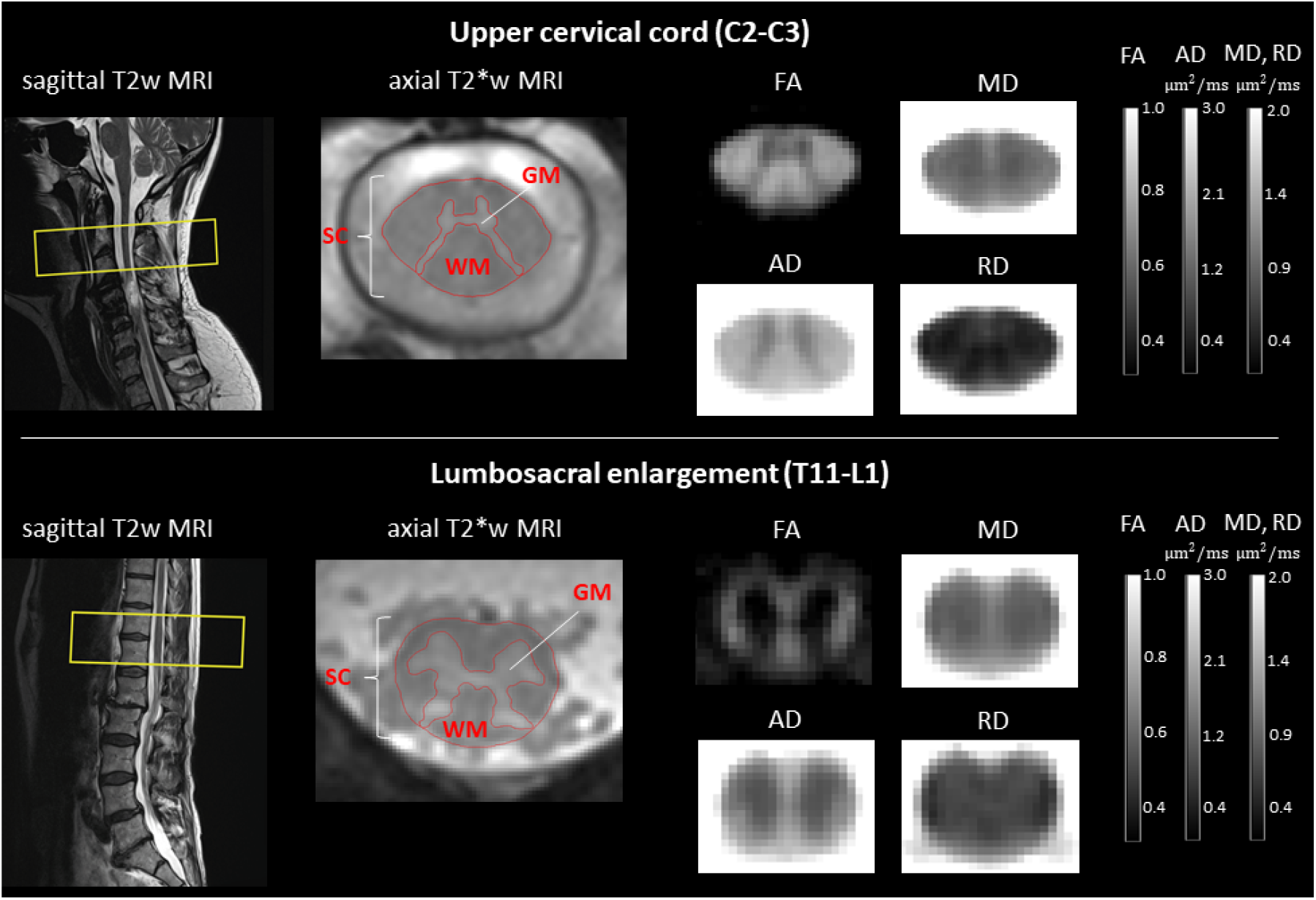
Examples of the acquired MRI scans and diffusion tensor imaging (DTI) maps, both in the upper cervical cord (C2-C3) and lumbosacral enlargement (T11-L1). T2*-weighted MRI (Siemens MEDIC) and diffusion MRI were acquired with identical slice prescription, defined on sagittal T2-weighted images with the slice stack (yellow rectangle) centered at the C2-C3 disc in the upper cervical cord and at the widest point of the spinal cord (SC) in the lumbosacral enlargement (usually between T11-L1). SC and gray matter (GM) were segmented manually on the T2*-weighted images to obtain cross-sectional areas. Notice the good contrast between GM and white matter (WM) and between WM and CSF. WM area was obtained by subtracting GM from the SC area. DTI maps including fractional anisotropy (FA), mean (MD), axial (AD), and radial diffusivity (RD) were computed from the diffusion MRI dataset. Shown is a single slice of the group-averaged FA, MD, AD, and RD maps. Notice the lower FA, MD, AD, but higher RD in the GM compared to the WM.

### Processing of diffusion-weighted images

Images were cropped to an in-plane FOV of 30×30 mm^2^, followed by slice-wise eddy-current and motion correction using the ECMOCO algorithm in ACID^37^. Maps of DTI metrics including fractional anisotropy, mean, axial, and radial diffusivity were obtained by fitting the tensor model on the corrected images using a robust tensor fitting algorithm^37,38^ (Fig. 1). The mean diffusion-weighted images were segmented for SC with *Propseg*^39^ and were normalized to the PAM50 template using Spinal Cord Toolbox^40^. The obtained deformation field was then applied to all DTI maps. Mean values of DTI metrics were extracted within the dorsal, lateral, and ventral WM columns. Binary masks of these WM columns were created by merging the probabilistic maps of 30 WM tracts included in the Spinal Cord Toolbox and thresholding them at 0.5.

### Processing of T2*-weighted images

The four MEDIC images were averaged using serial longitudinal registration in SPM12^41^ to account for between-volume motion. The average image was resliced to 5 mm thickness to increase signal-to-noise ratio. SC was segmented using the semi-automatic 3D active surface cord segmentation method as implemented in JIM 7.0^42^, while gray matter (GM) was segmented manually (Fig. 1). White matter (WM) mask was created by subtracting the GM from the SC mask. Cross-sectional tissue areas including SC, GM, and WM areas were extracted from the segmentations and were averaged across slices. In the lumbosacral enlargement, only three slices around the slice with the largest SC area were considered to ensure comparable and reproducible anatomical location.

### Clinical assessment

Neurological status and functional impairment were assessed according to the International Standards for Neurological Classification of Spinal Cord Injury (ISNCSCI) protocol.^43^ Single motor scores summed up between C5-T1 and L2-S1 yield the upper and lower extremity motor scores, respectively. Single light touch and pinprick scores summed up over across all neurological levels are referred to as total light touch and pinprick scores. DCM patients were assessed on the modified Japanese Orthopedic Scale (mJOA, max. 18 points).

### Statistical analysis

Differences in age and clinical scores between groups were assessed using pairwise Wilcoxon rank-sum tests, with the p-values corrected by the Benjamini-Hochberg procedure. One-way ANOVA was used to assess between-group differences in cross-sectional tissue areas (SC, GM, WM areas) and DTI metrics in the dorsal, lateral, and ventral WM columns. Significant differences between pairs of groups were tested using Tukey’s multiple comparison test for post-hoc analysis (p<0.05). Pearson correlation coefficient *r* quantified linear relationships between metrics measured at C2/C3 and in the lumbosacral enlargement. Images were carefully inspected and excluded if they were affected by extensive susceptibility and/or motion artifacts. As a result, one tSCI patient was excluded fully and another only from the analysis of cross-sectional areas in the lumbosacral enlargement. One healthy control was excluded from the DTI analysis at C2/C3.

## Results

### Patients’ characteristics and clinical outcomes

Demographic and clinical information is listed in Table 1 for DCM and Table 2 for tSCI patients. In the DCM cohort, the maximal cervical spinal stenosis was at C4/C5 in 2, C5/C6 in 14, and C6/C7 in 2 patients. Of the 18 patients, 10 had multi-segmental cervical spinal canal stenosis. 14 DCM patients had mild (mJOA≥15) and 4 patients moderate (12≤mJOA≤14) DCM. In the tSCI cohort, 1 patient was AIS C, 12 AIS D, and 1 AIS E (Table 2). There was no significant age difference between tSCI patients and controls (p=0.213) and between DCM and tSCI patients (p=0.126), but DCM patients were older than controls (p=0.034). Compared to DCM patients, tSCI patients had lower extremity motor score (tSCI vs. DCM, mean (±std), 43.5 (±11.1) vs. 50 (±0), p=0.002), but not upper extremity motor score (48.3 (±2.6) vs. 49.6 (±1.2), p=0.131), pinprick (91.1 (±24.6) vs. 105.5 (±9.1), p=0.063), and light touch scores (98.6 (±15.8) vs. 105.9 (±9.8), p=0.190).

**Table 2.** Demographic and clinical Information of the tSCI patients.

| ID | Sex | Age (yrs) | Time since injury [m] | Neurological injury level | AIS | UEMS score | LEMS score | LT score | PP score |
| --- | --- | --- | --- | --- | --- | --- | --- | --- | --- |
| 1 | M | 41-45 | 12.2 | C6 | D | 50 | 50 | 111 | 112 |
| 2 | M | 66-70 | 13.2 | C7 | D | 49 | 44 | 110 | 107 |
| 3 | M | 46-50 | 15.1 | C8 | D | 50 | 50 | 94 | 103 |
| 4 | M | 26-30 | 4.9 | C7 | D | 46 | 23 | 74 | 45 |
| 5 | M | 41-45 | 4.6 | C6 | D | 45 | 44 | 90 | 84 |
| 6 | M | 46-50 | 1.8 | C5 | D | 50 | 50 | 112 | 107 |
| 7 | M | 26-30 | 11.3 | C7 | C | 50 | 14 | 70 | 44 |
| 8 | M | 61-65 | 4.1 | C2 | D | 44 | 45 | 79 | 64 |
| 9 | M | 56-60 | 2.7 | C3 | D | 43 | 45 | 87 | 75 |
| 10 | M | 51-55 | 2.4 | C5 | D | 50 | 50 | 112 | 105 |
| 11 | M | 51-55 | 2.1 | C4 | E | 50 | 50 | 112 | 112 |
| 12 | M | 61-65 | 1.3 | T1 | D | 50 | 50 | 112 | 112 |
| 13 | M | 36-40 | 1.1 | C7 | D | 50 | 50 | 110 | 97 |
| 14 | M | 36-40 | 3.3 | T1 | D | 49 | 44 | 108 | 108 |

AIS = American Spinal Injury Association Impairment Scale; UEMS = International Standards for Neurologic Classification of Spinal Cord Injury (ISNCSCI) upper extremity motor score (max. 50); LEMS = ISNCSCI lower extremity motor score (max. 50); LT = ISNCSCI light touch score (max. 112); PP = ISNCSCI pinprick score (max. 112).

### Diffusion tensor imaging metrics (microstructure)

In the lumbosacral enlargement, DCM patients did not show significant differences in any DTI metric compared to controls (Table 3, Fig. 2). tSCI patients, on the other hand, had lower fractional anisotropy in the lateral (tSCI vs. controls, −15.5%, p<0.001) and ventral WM columns (−10.4%, p<0.001) and showed strong trend toward lower values in the dorsal column (−9.0%, p=0.057). Radial diffusivity was higher in the lateral column (tSCI vs. controls, +13.2%, p=0.018). Compared to DCM patients, tSCI patients had lower fractional anisotropy in the lateral (tSCI vs. DCM, −11.9%, p=0.007) and ventral column (−8.0%, p=0.021), and showed trend toward lower values in the dorsal column (−8.9%, p=0.068).

**Table 3.** List of tissue-specific cross-sectional areas and column-specific diffusion tensor imaging (DTI) metrics. Values represent mean and standard deviation across participants, separately for each region (C2/C3 and LSE) and group (healthy controls, degenerative cervical myelopathy (DCM) patients, and traumatic spinal cord injury (tSCI) patients).

|  |  |  | ROI | controls | DCM | SCI | DCM - controls |  | tSCI - controls |  | tSCI - DCM |  |
| --- | --- | --- | --- | --- | --- | --- | --- | --- | --- | --- | --- | --- |
|  |  |  |  |  |  |  | difference | p-value | difference | p-value | difference | p-value |
| Cross-sectional areas<br>(mm<sup>2</sup>) | C2/C3 | SC |  | 89.4 ± 8.0 | 76.5 ± 7.1 | 73.7 ± 8.9 | -14.4% | **<0.001** | -17.6% | **<0.001** | -3.7% | 0.598 |
|  |  | WM |  | 76.1 ± 7.5 | 63.9 ± 6.3 | 62.3 ± 7.6 | -16.0% | **<0.001** | -18.1% | **<0.001** | -2.5% | 0.814 |
|  |  | GM |  | 13.3 ± 0.9 | 12.6 ± 1.3 | 11.4 ± 1.8 | -5.0% | 0.270 | -14.2% | **<0.001** | -9.7% | **0.035** |
|  | LSE | SC |  | 69.2 ± 7.1 | 59.3 ± 5.6 | 62.4 ± 7.5 | -14.4% | **<0.001** | -9.9% | **0.017** | 5.2% | 0.423 |
|  |  | WM |  | 48.1 ± 5.8 | 40.0 ± 5.3 | 43.0 ± 5.7 | -16.9% | **<0.001** | -10.5% | **0.043** | 7.7% | 0.313 |
|  |  | GM |  | 21.1 ± 2.6 | 19.3 ± 1.4 | 19.5 ± 2.8 | -8.7% | **0.045** | -7.7% | 0.142 | 1.1% | 0.964 |
| FA | C2/C3 | WM dor |  | 0.68 ± 0.05 | 0.63 ± 0.05 | 0.61 ± 0.08 | -7.9% | **0.024** | -10.0% | **0.007** | -2.3% | 0.781 |
|  |  | WM lat |  | 0.70 ± 0.04 | 0.66 ± 0.04 | 0.61 ± 0.08 | -6.2% | **0.039** | -13.3% | **<0.001** | -7.6% | **0.029** |
|  |  | WM ven |  | 0.61 ± 0.05 | 0.58 ± 0.04 | 0.56 ± 0.05 | -4.7% | 0.197 | -8.2% | **0.023** | -3.7% | 0.471 |
|  | LSE | WM dor |  | 0.54 ± 0.05 | 0.54 ± 0.06 | 0.49 ± 0.06 | -0.1% | 0.999 | -9.0% | 0.057 | -8.9% | 0.068 |
|  |  | WM lat |  | 0.53 ± 0.05 | 0.51 ± 0.06 | 0.45 ± 0.04 | -4.1% | 0.407 | -15.5% | **<0.001** | -11.9% | **0.007** |
|  |  | WM ven |  | 0.49 ± 0.04 | 0.48 ± 0.04 | 0.44 ± 0.03 | -2.5% | 0.584 | -10.4% | **<0.001** | -8.0% | **0.021** |
| MD<br>(μm<sup>2</sup>/ms) | C2/C3 | WM dor |  | 1.17 ± 0.12 | 1.28 ± 0.13 | 1.23 ± 0.19 | 9.3% | 0.064 | 4.9% | 0.513 | -4.0% | 0.588 |
|  |  | WM lat |  | 1.10 ± 0.08 | 1.16 ± 0.12 | 1.18 ± 0.13 | 5.2% | 0.255 | 7.2% | 0.116 | 1.9% | 0.841 |
|  |  | WM ven |  | 1.17 ± 0.14 | 1.19 ± 0.14 | 1.28 ± 0.17 | 1.8% | 0.905 | 9.7% | 0.111 | 7.8% | 0.226 |
|  | LSE | WM dor |  | 1.22 ± 0.11 | 1.18 ± 0.13 | 1.21 ± 0.16 | -3.3% | 0.627 | -0.6% | 0.985 | 2.7% | 0.785 |
|  |  | WM lat |  | 1.00 ± 0.11 | 0.99 ± 0.09 | 1.03 ± 0.13 | -0.9% | 0.966 | 3.2% | 0.695 | 4.1% | 0.566 |
|  |  | WM ven |  | 1.16 ± 0.12 | 1.20 ± 0.09 | 1.21 ± 0.14 | 3.7% | 0.552 | 5.0% | 0.374 | 1.2% | 0.942 |
| AD<br>(μm<sup>2</sup>/ms) | C2/C3 | WM dor |  | 2.23 ± 0.13 | 2.28 ± 0.16 | 2.18 ± 0.21 | 2.1% | 0.662 | -2.2% | 0.675 | -4.2% | 0.245 |
|  |  | WM lat |  | 2.14 ± 0.09 | 2.13 ± 0.15 | 2.08 ± 0.09 | -0.7% | 0.927 | -2.9% | 0.294 | -2.3% | 0.481 |
|  |  | WM ven |  | 2.04 ± 0.17 | 2.02 ± 0.20 | 2.12 ± 0.18 | -0.9% | 0.952 | 4.2% | 0.430 | 5.1% | 0.292 |
|  | LSE | WM dor |  | 2.05 ± 0.17 | 1.97 ± 0.18 | 1.94 ± 0.19 | -4.0% | 0.339 | -5.4% | 0.194 | -1.5% | 0.893 |
|  |  | WM lat |  | 1.67 ± 0.19 | 1.62 ± 0.17 | 1.59 ± 0.23 | -2.9% | 0.720 | -4.8% | 0.490 | -1.9% | 0.903 |
|  |  | WM ven |  | 1.85 ± 0.19 | 1.89 ± 0.13 | 1.85 ± 0.19 | 2.2% | 0.774 | 0.3% | 0.997 | -1.9% | 0.851 |
| RD<br>(μm<sup>2</sup>/ms) | C2/C3 | WM dor |  | 0.64 ± 0.13 | 0.78 ± 0.14 | 0.75 ± 0.20 | 22.0% | **0.020** | 17.4% | 0.118 | -3.8% | 0.858 |
|  |  | WM lat |  | 0.58 ± 0.08 | 0.68 ± 0.12 | 0.73 ± 0.17 | 15.9% | 0.062 | 25.7% | **0.003** | 8.5% | 0.400 |
|  |  | WM ven |  | 0.73 ± 0.14 | 0.77 ± 0.12 | 0.86 ± 0.17 | 5.6% | 0.666 | 17.4% | 0.050 | 11.2% | 0.238 |
|  | LSE | WM dor |  | 0.80 ± 0.11 | 0.78 ± 0.13 | 0.85 ± 0.17 | -2.4% | 0.899 | 5.4% | 0.634 | 8.0% | 0.408 |
|  |  | WM lat |  | 0.66 ± 0.09 | 0.68 ± 0.08 | 0.75 ± 0.09 | 1.7% | 0.916 | 13.2% | **0.018** | 11.3% | **0.049** |
|  |  | WM ven |  | 0.81 ± 0.10 | 0.85 ± 0.09 | 0.89 ± 0.12 | 5.4% | 0.438 | 10.3% | 0.069 | 4.7% | 0.564 |

AD = axial diffusivity; DCM = degenerative cervical myelopathy; FA = fractional anisotropy; GM = gray matter; LSE = lumbosacral enlargement; RD = radial diffusivity; tSCI = traumatic spinal cord injury; WM = white matter; WM dor = dorsal white matter column; WM lat = lateral white matter column; WM ven = ventral white matter column.

**Fig. 2.**
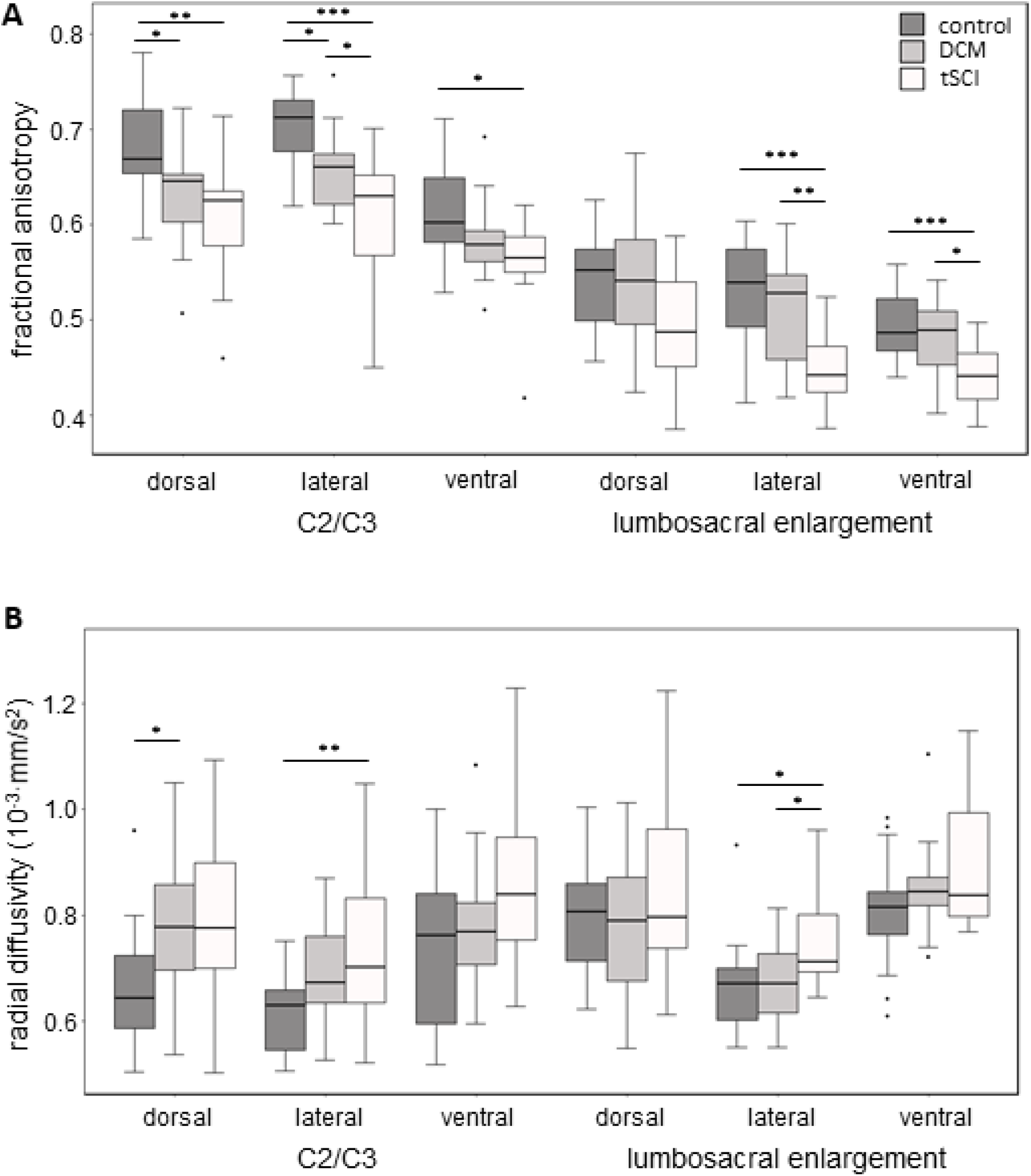
Boxplot of fractional anisotropy (A) and radial diffusivity (B) values in healthy controls, DCM patients, and tSCI patients. Values are plot separately for the dorsal, lateral, ventral column, and for the C2/C3 and lumbosacral enlargement. P-values less than 0.05, 0.01, and 0.001 are designated with *, **, and ***, respectively.

At C2/C3, compared to controls, DCM patients showed decreased fractional anisotropy in the dorsal (DCM vs. controls, −7.9%, p=0.024) and lateral column (−6.2%, p=0.039) and increased radial diffusivity (+22.0%, p=0.020) in the dorsal column (Table 3, Fig. 2). tSCI patients had lower fractional anisotropy in all WM columns (tSCI vs. controls; dorsal: −10.0%, p=0.007; lateral: −13.3%, p<0.001; ventral: −8.2%, p=0.023) and higher radial diffusivity in the lateral (+25.7%, p=0.003) and ventral column (+17.4%, p=0.050). Compared to DCM patients, tSCI did not have significantly different DTI metrics.

### Cross-sectional tissue areas (macrostructure)

In the lumbosacral enlargement, compared to controls, DCM patients had smaller SC (DCM vs. controls: −14.4%, p<0.001), WM (−16.9%, p<0.001), and GM area (−8.7%, p=0.045) than controls (Table 3, Fig. 3). Similarly, tSCI patients had smaller SC area (tSCI vs. controls: −9.9%, p=0.017) and WM area (−10.5%, p=0.043). No statistical differences between tSCI and DCM were detectable.

**Fig. 3.**
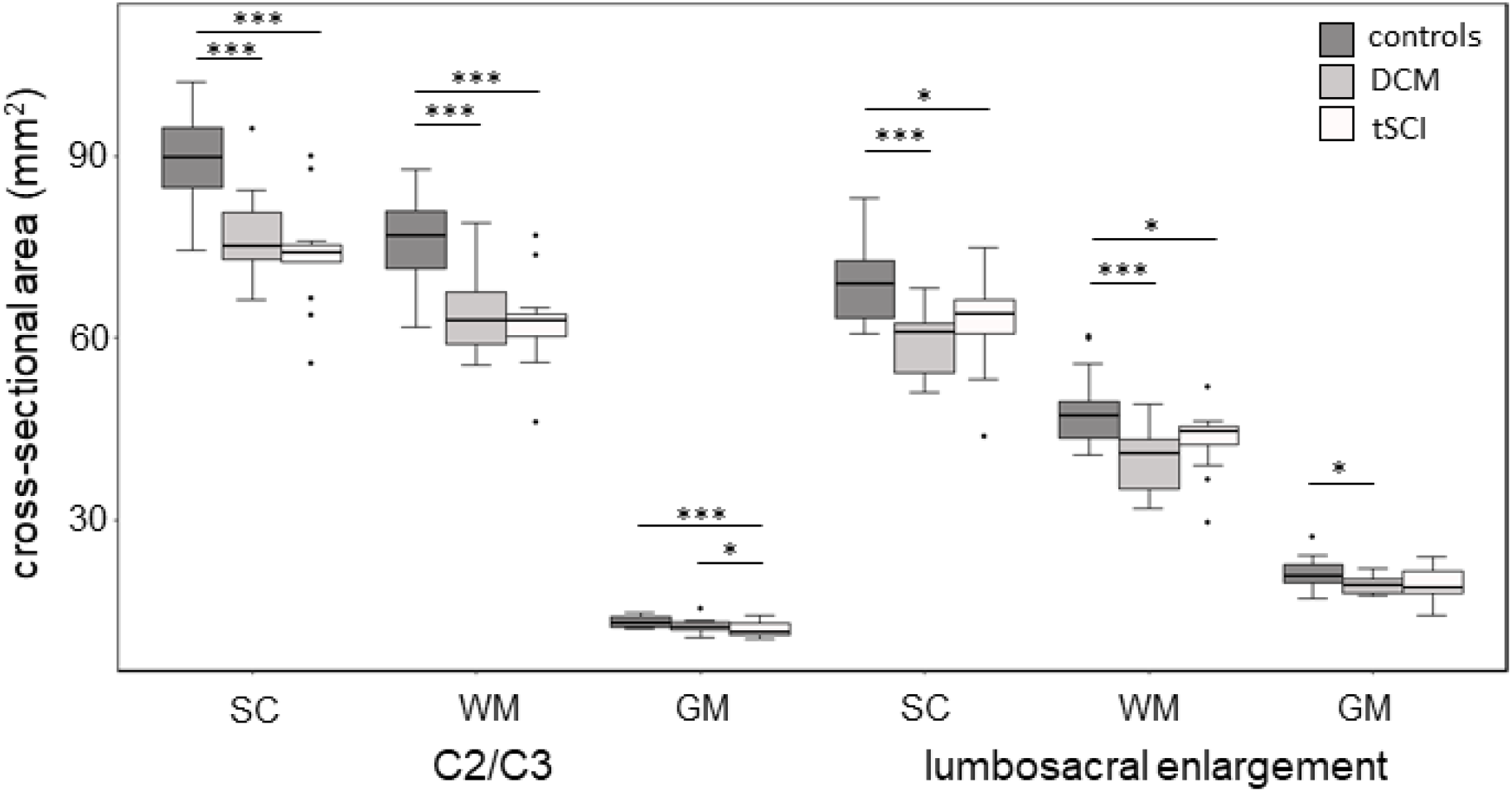
Boxplot of cross-sectional area of spinal cord (SC), white matter (WM), and gray matter (GM) in healthy controls, DCM patients, and tSCI patients. Values are plot separately for C2/C3 and the lumbosacral enlargement. P-values less than 0.05, 0.01, and 0.001 are designated with *, **, and ***, respectively.

At C2/C3, compared to controls, DCM patients had smaller SC (DCM vs. controls: −14.4%, p<0.001) and WM (−16.0%, p<0.001) area, while tSCI patients had smaller SC (tSCI vs. controls: −17.6%, p<0.001), WM (−18.1%, p<0.001) and GM area (−14.2%, p<0.001) (Table 3, Fig. 3). Compared to DCM patients, tSCI patients had smaller GM area (tSCI vs. DCM: - 9.7%, p=0.035).

## Discussion

This study highlights disease-specific differences across the spinal cord axis, caudal and rostral to the primary injury. Compared to DCM patients, tSCI patients exhibited more severe microstructural changes in the WM of the lumbosacral enlargement. At C2/C3 (rostral to stenosis/injury), no differences were observed in the magnitude of WM degeneration between DCM and tSCI patients. Interestingly, both patient groups seemed to exhibit similar degree of tissue atrophy in the lumbosacral enlargement despite tSCI patients having more severe microstructural damage.

### Diffusion tensor imaging metrics (microstructure)

Beside the primary injury, an insult to the spinal cord also leads to widespread and distant changes along the whole neuroaxis via secondary axonal degenerative processes, resulting in irreversible damage to the myelin sheath and the axonal cytoarchitecture.^6,15,44^ Remote degenerations, both rostral and caudal to the stenosis/injury, of the WM and GM were previously demonstrated in vivo after DCM^45–47^ and cervical tSCI^26–28,48^. When comparing mild to moderate DCM to incomplete tSCI patients, we add to this by showing that tSCI patients exhibit more severe microstructural degenerative changes in the WM of the lumbosacral enlargement. In fact, DTI metrics in DCM patients were not significantly different to those in healthy controls, while tSCI patients had 9-16% lower fractional anisotropy and 5-13% higher radial diffusivity values (also shown in David et al., 2019^28^). Radial diffusivity has been linked to myelin content and integrity in various dys- and demyelination models^49–53^, while a decrease in fractional anisotropy can indicate both demyelination and axonal injury.^50,54–56^ This suggests that demyelination is likely to be the dominant degeneration process in the lumbosacral enlargement of incomplete tSCI patients. In contrast to our findings, a previous study found increased mean, axial, and radial diffusivity in the lumbosacral cord of DCM^34^, which might be due to the different cohorts and type of analysis (template-based column-specific analysis vs. whole WM analysis).

The reason behind tSCI patients having more altered DTI values in the lumbosacral enlargement is not clear; a potential explanation is that compensatory mechanisms might limit the overall amount of microstructural damage in DCM owing to the slowly progressing nature of the condition (see Figure 4 for potential mechanisms). tSCI, on the other hand, represents a blunt insult interrupting the axons, followed by rapid degeneration along the neuroaxis,^54^ which leaves less opportunity for compensatory mechanisms. By 2 months post-injury, significant WM damage has already been shown in the lateral column of the lumbosacral enlargement with fractional anisotropy being 10% lower than in healthy controls.^57^ Another explanation can be that the microstructural changes associated with DCM are more local with degenerative changes lagging at larger distances. This notion is supported by the findings at C2/C3 located only a few segments rostral to the injury level in contrast to the lumbosacral enlargement which is in at least 12 segments caudal.

**Fig. 4.**
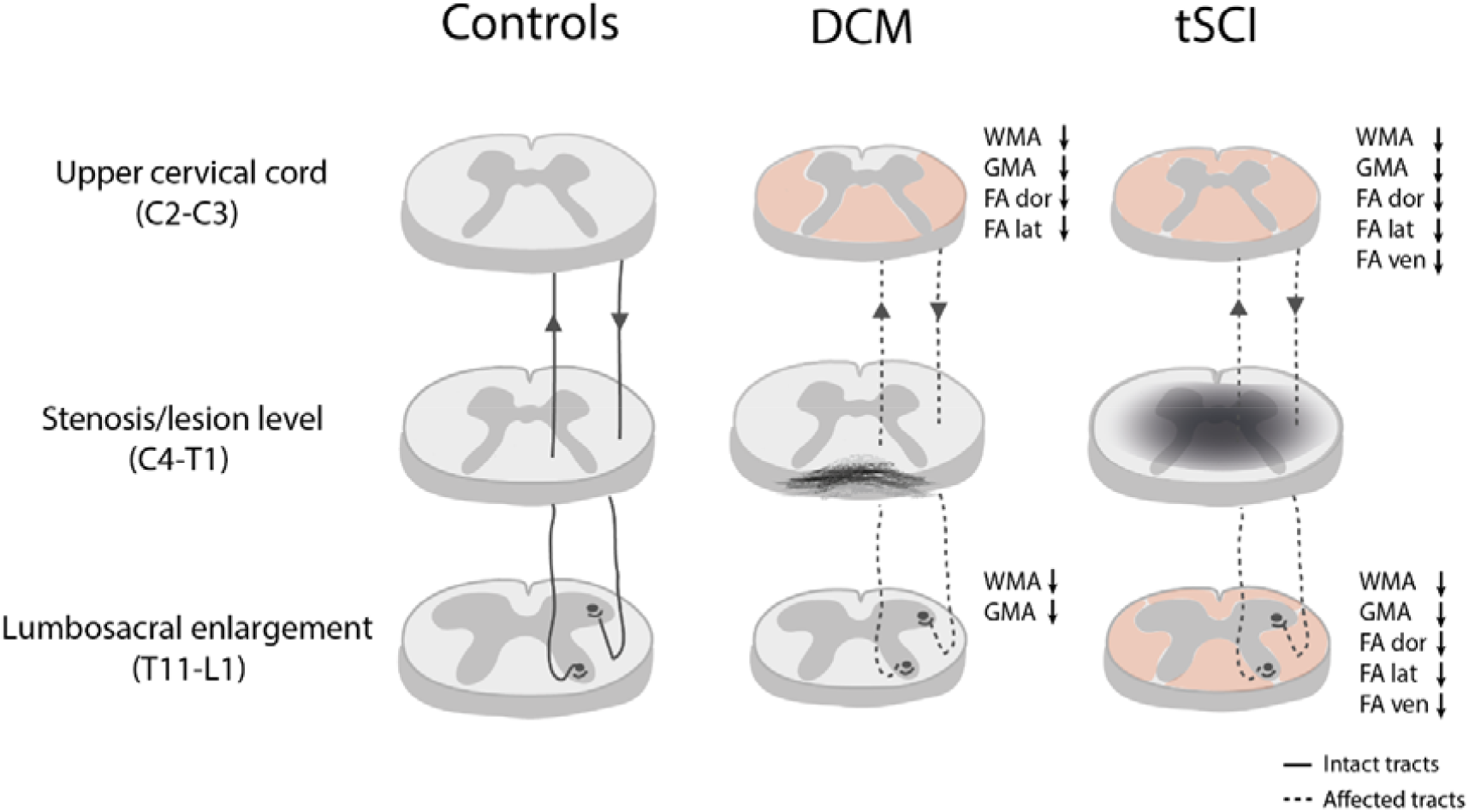
Visual summary of the findings from the analysis of cross-sectional tissue areas and DTI maps in degenerative cervical myelopathy (DCM) and traumatic spinal cord injury (tSCI) patients with incomplete cervical injuries. Significantly lower fractional anisotropy (FA) within a white mater (WM) column, compared to controls, is indicated by red shading. FA was selected for visualization because this DTI metric showed the most changes relative to controls. In addition, the size of the schematic slices in the upper cervical cord and lumbosacral enlargement is scaled proportionally to the measured spinal cord area, compared to the corresponding slice in controls. While the primary injury mechanisms in degenerative cervical myelopathy (DCM) and traumatic spinal cord injury (tSCI) are fundamentally different, with tSCI being a sudden traumatic event and DCM resulting from progressive degeneration of the cervical spine, they share similar secondary injury mechanisms involving axonal degeneration and accompanying demyelination.^2,6^ This neurodegeneration propagates rostral and caudal to the primary injury site (dotted lines) in both DCM and tSCI, via anterograde, retrograde, and trans-synaptic degenerations. As a result, tSCI patients showed FA decrease in the dorsal, lateral, and ventral WM columns both rostral (upper cervical cord) and caudal (lumbosacral enlargement) to the incomplete cervical injury. In contrast, DCM patients were found to have decreased FA only rostral to the lesion, in the dorsal and lateral WM columns. Interestingly, the observed WM and gray matter decreases, relative to controls, were comparable in DCM and tSCI at both levels.

At C2/C3, DCM patients exhibited altered fractional anisotropy and radial diffusivity compared to healthy controls, the magnitude of which was similar to those in tSCI patients as demonstrated previously^29^. However, the spatial distribution of these changes was not uniform; while our and previous results^45,58^ indicate strong secondary WM damage in the dorsal and lateral column rostral to stenosis, the differences were found to be negligible in the ventral horn. In contrast, and confirming previous findings^27,48,59^, incomplete tSCI resulted in severe and widespread WM damage across all WM columns at C2/C3. Previous histopathological analyses of DCM showed evidence of both anterograde and retrograde degeneration of sensory tracts in the dorsal column^60,61^ and motor tracts in the lateral column^61–63^, with the former being linked to poor or disturbed sensation and the latter to spastic gait, but less axonal damage in the ventral column^64^.

### Cross-sectional tissue areas (macrostructure)

Remote atrophy of neural tissue occurs as a result of accumulation of degenerative changes, usually leading to irreversible tissue loss. At C2/C3, we confirmed a previous observation^29^ and found no difference in SC and WM area between DCM and incomplete tSCI patients. In the lumbosacral enlargement, DCM and tSCI patients had similar SC and WM area of ca. 10-17% smaller than in healthy controls, shown previously for DCM^34^ and tSCI patients^28^. In both patient groups, GM area was around 7-9% smaller than in healthy controls, demonstrating similar degree of remote GM atrophy in the lumbosacral enlargement. GM atrophy caudal to the injury is well known after tSCI, affecting e.g. the lower motor neurons.^65,66^ Histopathology of chronic compression in DCM has also revealed degeneration of central GM, occurring typically in patients with poor sensation, proprioception defects or sphincter disturbance^60^, which represented 61% of our DCM patients. Rostral to the injury, tSCI patients showed more GM atrophy than DCM patients, which might be explained by the differential pathophysiology above and below the lesion. While GM atrophy below the lesion occurs primarily via trans-synaptic degeneration^67^, above the lesion it is thought to occur primarily via the perturbation of propriospinal network^68^, which seems to be more pronounced in tSCI patients.

Atrophy in the lumbosacral cord of DCM patients without concomitant DTI alterations is interesting. On one hand, it might indicate a mere compression of the tissue, consequence a chronic compression of the cervical cord, without measurable microstructural changes. On the other hand, it might be related to the unspecific nature of cross-sectional areas, as tissue atrophy can be the result of several pathophysiological processes, some of which DTI might not be sensitive to. Clearly, the exact contribution of pathophysiological mechanisms to the atrophy is not fully understood and warrants further research.

### Limitations

There were demographic and clinical differences between DCM and tSCI patients. tSCI patients had lower ISNCSCI lower extremity motor scores, which might contribute to the more altered DTI values in the lateral and ventral columns of the lumbosacral enlargement. However, the fact that strong group differences were observed even in the dorsal column despite similar light touch scores implies that the observed findings represent disease-specific and not severity-specific differences. Also, DCM patients were on average 7.9 years older than tSCI patients. While the cord atrophy rate due to normal aging is estimated to be 0.06% per year^69^, this would account for a group difference of only 0.5%, in contrast to the observed group differences of 10-15%. Since the onset of symptoms in DCM patients is not known, there might be a considerable variability in the stage of progression in which participants entered the study. Patients with a later stage of progression were exposed longer to chronic compression but also had more time for compensatory mechanisms.

## Conclusion

This study suggests that mild to moderate DCM leads to anterograde degeneration of the dorsal columns and retrograde degeneration in the lateral column at C2/C3 (rostral to stenosis), but only to minimal changes in the lumbosacral enlargement (caudal to stenosis). Incomplete tSCI leads to more widespread WM damage, affecting the whole WM both in the lumbosacral enlargement and at C2/C3. These remote degenerative changes are important because they are likely to contribute to the patients’ impairment and recovery potential. Therefore, clinical trials should target remote degeneration before irreversible damage (atrophy) occurs, e.g. by means of neuroprotective agents. Advanced MRI techniques, such as diffusion MRI are sensitive tools to assess remote pathological changes in both DCM and tSCI patients.

## Data Availability

The authors certify they have documented all data, methods,
and materials used to conduct the research presented. Anonymized data pertaining to the research presented will be
made available by request from qualified investigators.

## Acknowledgments

We would like to thank all the patients and healthy volunteers who participated in this study. We also thank the staff of the Department of Radiology and Neurology at the University Hospital Balgrist.

